# Proxalutamide (GT0918) Reduces the Rate of Hospitalization in mild-to-moderate COVID-19 Female Patients: A Randomized Double-Blinded Placebo-Controlled Two-Arm Parallel Trial

**DOI:** 10.1101/2021.07.06.21260086

**Authors:** Flávio Adsuara Cadegiani, Ricardo Ariel Zimerman, Daniel do Nascimento Fonseca, Michael do Nascimento Correia, John McCoy, Carlos Gustavo Wambier, Andy Goren

**Author notes:** **Corresponding author**: Flavio Adsuara Cadegiani, MD, PhD, Applied Biology, Inc., 17780 Fitch, Irvine, CA 92614.

## Abstract

**Background:** Antiandrogens were shown to be effective in mild-to-moderate COVID-19 male patients, supported by the SARS-CoV-2 dependency on transmembrane protease, serine 2 (TMPRSS2), which is solely modulated by androgens, for cell entry. While women with hyperandrogenism experiment more symptoms in COVID-19 compared to women without hyperandrogenism, and the chronic use of an antiandrogen seemed to mitigate these symptoms, whether the benefits would be observed in overall females is unknown. The objective of this study is to evaluate the efficacy of proxalutamide as a treatment for mild-to-moderate COVID-19 in females.

**Methods:** This was a randomized, double-blind, placebo-controlled, two-arm, parallel study on COVID-19 female outpatients, that compared the use of proxalutamide versus placebo. The primary outcome was hospitalization rates throughout 30 days after randomization. Patients with laboratory confirmed COVID-19 not hospitalized were recruited in two sites in Brasilia, Brazil, between January 4 and February 28, 2021, were randomized on a 2:3 ratio between proxalutamide and placebo, and were administered proxalutamide 200mg/day or placebo for seven days, in addition to usual care.

**Results:** A total of 177 women were randomized, including 75 in the proxalutamide arm and 102 patients in the placebo arm. None of the patients lost follow-up or discontinued treatment. The 30-day hospitalization rate was 2.7% in the proxalutamide arm and 18.6% in the placebo arm (p<0.001), with a hospitalization risk ratio (RR) of 0.14 [95% confidence interval (CI), 0.03-0.59].

**Conclusions:** These findings suggest that treatment of COVID-19 patients with proxalutamide in combination with standard of care was reduced hospitalization rate by 86% (p < 0.001), with no safety concerns. (Clinicaltrials.gov: NCT04853134)

## Introduction

During the COVID-19 pandemic, it has been extensively demonstrated that men are more likely to be infected, more likely to have severe disease, and have a greater case fatality rate compared to women.^1,2^

Although lifestyle differences and gender-biased comorbidities may have contributed to this gender discrepancy, these factors unlikely explain the overwhelming differences. Alternatively, it has been suggested that the male bias in COVID-19 disease severity may be linked to androgens.^3,4^

For the cell entry, SARS-CoV-2 spike protein needs to be primed by the transmembrane protease, serine 2 (TMPRSS2), that is expressed on the surface of human cells.5 The only known promoters of the TMPRSS2 gene and TMPRSS2 expression are androgens.^6^ Correspondingly, several observation studies, the androgen-mediated phenotype of androgenetic alopecia (AGA) as well as differences in sensitivity of the androgen receptor (AR) have been linked to COVID-19 disease severity.^3,4,7-9^

It would follow that reducing the expression of TMPRSS2 by blocking the androgen receptor would decrease SARS-CoV-2 entry into human cells. Indeed, evidence connecting COVID-19 to antiandrogens have emerged from several sources.^10-14^

Although initially it has been considered that androgens would play a relevant role in COVID-19 only in males, females with hyperandrogenism, that could theoretically be under higher risk of severe COVID-19, were indeed correlated with more symptoms and more severity.^15-17^ In accordance, preliminary observational data showed that women with hyperandrogenism that used antiandrogens not only had the increased risk mitigated, but also had fewer symptoms when compared to non-hyperandrogenic women.^18^

Taken together, there is growing body of evidence to support that SARS-CoV-2 infectivity is mediated by the androgen receptor and will likely respond to drugs that reduce androgen receptor function. Two major drug classes that are promising agents against COVID-19. Fist, the 5-alpha-reductase inhibitors (5ARis), that are commonly prescribed for androgenetic alopecia and benign prostatic hyperplasia, and they act by blocking the conversion of testosterone to a 5-times more potent androgen, dihydrotestosterone (DHT).^10,13,14,19^ Second, the non-steroidal antiandrogens (NSAA), used for castrated-resistant prostate cancer.^11,12^

Proxalutamide (GT0918) is a novel second generation NSAA that has demonstrated several distinct advantages as a therapy for SARS-CoV-2. Proxalutamide demonstrates at least three mechanisms of action potentially protective against COVID-19: antagonism of TMPRSS2 expression, regulator of the angiotensin converting enzyme-2 (ACE2), and as a potent anti-inflammatory agent.

The inhibition of TMPRSS2 expression occurs through its highly effective ability to block AR, mediated by two pathways: acting as an antagonist of AR and as a down-regulator of AR expression. It has also been reported that proxalutamide lowers the expression of ACE2, an expected added benefit for preventing SARS-CoV-2 entry into lung cells. Finally, proxalutamide demonstrated in vitro ability to reduce tumor necrosis factor-alpha (TNF-alpha) and other inflammatory cytokines substantially.^20^

The downregulation of AR expression, the regulation of ACE2 and the anti-inflammatory property are mechanisms that have not been described in other NSAA, for example, bicalutamide or enzalutamide. Because of these multiple mechanisms of action, it is expected to be a more effective and less toxic second-generation anti-androgen drug therapy.

Because of the abilities other than those attributed to androgens and the fact that proxalutamide acts in the AR receptor, that tends to be upregulated in women due to lower androgen circulating levels, we hypothesized that proxalutamide could be effective not only in men, but also in women. In terms of safety profile for its use in women, it has been studied and certified in both men and women. The objective of this study is to evaluate the efficacy of proxalutamide as a treatment for mild-to-moderate COVID-19 in females.

## Methods

This was a randomized, double-blind, placebo-controlled study of proxalutamide compared with placebo in subjects with laboratory confirmed primary SARS-CoV-2 infection, aimed to demonstrate the superiority of proxalutamide over placebo in reducing the rate of COVID-19 hospitalizations within 30 days after initiation of treatment. The planned total sample size was approximately 168 subjects randomized in a 1:2 ratio (Appendix 1).

This study was conducted in accordance with the ethical principles that have their origin in the Declaration of Helsinki and that are consistent with Good Clinical Practices and applicable regulatory requirements. Known instances of nonconformance were documented and are not considered to have had an impact on the overall conclusions of this study. The study was approved to be conducted in females by the Brazilian National Ethics Committee of the Ministry of Health [Comitê de Ética em Pesquisa (CEP) da Comissão Nacional de Ética em Pesquisa (CONEP) do Ministério da Saúde (MS) (CEP/CONEP/MS]. Approval number for females: #4.513.428; process number (CAAE): 36703320.8.0000.0023. Registration number in Clinicaltrials.gov: NCT04853134. All patients were volunteers and received no financial compensation. All patients gave informed consent.

### Subject selection

Subjects were recruited through the clinic e-mail list, social media, referred by primary care physicians, or by emergency units that did not detect signs of severity, in known reverse transcription Polymerase Chain Reaction (rtPCR) positive COVID-19 patients. Self-referred patients underwent medical screening and rtPCR SARS-CoV-2 (Automatized Platform, Cobas, Roche, USA) collected using nasopharyngeal swabs, in case of non-confirmed cases, prior to enrollment and randomization, between January 4 and February 28, 2021.

Subjects enrolled in this study were required to meet the following key acceptance criteria: females aged 18 years old or above, with mild-to-moderate COVID-19 (COVID-19 8-point ordinary scale < 3), confirmed for COVID-19 through positive rtPCR-SARS-CoV-2 in the past seven days, not hospitalized for acute respiratory symptoms, with normal kidney function (Serum creatinine ≤ 1.5xULN or creatinine clearance ≥ 60 mL/min, calculated using Cockcroft-Gault formula, enzymatic assay), absence of liver injury (ALT < 150 U/L or AST < 120 U/L; IFCC without pyridoxal phosphate assay), with normal coagulation (INR ≤ 1.5 ULN, and APTT ≤ 1.5 ULN), not pregnant or willing to be pregnant in the three weeks after treatment, with effective contraceptive methods (if applicable), not breastfeeding, with written informed consent obtained prior to any screening procedures.

Subjects were not to be enrolled into the study if it was determined upon pre-study examination that they met the following key criteria: enrolled in a study to investigate another COVID-19 drug or any experimental drug in the past four months, taking an anti-androgen of any type including, known to be allergic to the investigational product or similar drugs or any excipients, with malignant tumors in the past five years (except for carcinoma in situ of any type completely resected), with known severe cardiovascular diseases including myocardial infarction in the past six months or arterial thrombosis, congestive heart failure class three or four according to the New York Heart Association (NYHA) classification, uncontrolled medical conditions that could compromise, such as uncontrolled hypertension, diabetes mellitus or autoimmune diseases, known diagnosis of human immunodeficiency virus (HIV), hepatitis C, active hepatitis B, or treponema pallidum (testing is not mandatory), and if not willing or unable to provide informed consent.

Patients informed baseline characteristics, comorbidities, chronic use of medications and previous test results. Active search was performed for type 2 diabetes mellitus (T2DM), hypertension, obesity as defined as body mass index (BMI) above 30 kg/m^2^, and chronic obstructive pulmonary disease (COPD).

### Randomization

Eligible patients were be randomized to receive either proxalutamide (arm 1) 200mg/day (2 tablets of 100mg daily) for seven days or placebo (arm 2) (2 tablets of placebo daily). Proxalutamide 100mg and placebo were manufactured to have identical characteristics (Kintor Pharmaceuticals Ltd., Suzhou, China).

To avoid repetition of the randomization process of those performed for male patients,^21^ a different blocked randomization list was created with block size of 25 instead of 10 and the sequence of dispensing was followed throughout the trial, in large 25-size blocks per arm. Each subject was assigned a subject study number. The first subject was assigned the number 001 and each subject thereafter will be assigned a consecutive number i.e., 002, 003, etc. The randomization plan was based on a 1:2 ratio between active (Arm 1) and placebo (Arm 2) arms due to the uncertain safety profile of proxalutamide in COVID-19 on females. Due to the high efficacy of proxalutamide observed in COVID-19 male outpatients,^21^ the distribution of the blocks different from typically expected sizes. Since the study was double-blinded, the following randomization schedule was used but the identification of the arm assignment was only known by the study monitor: Subjects 001-025 were assigned to Arm 1; Subjects 026-050 were assigned to Arm 2; Subjects 051-075 were assigned to Arm 1; Subjects 076-100 were assigned to Arm 2; Subjects 101-125 were assigned to Arm 1; Subjects 126-150 were assigned to Arm 2; Subjects 151-175 were assigned to Arm 1; Subjects 176-200 were assigned to Arm 2; Subjects 201-225 were assigned to Arm 1; etc. This sequence of blocks of 25 repeated continuously. The group assignment was known only to the study monitor. The patients and doctors directly managing patient care were blinded to the study group allocation until the close of the study. Patients were instructed to take one tablet once a day. The telephone contact and clinical follow-up were independently performed by study assistants that were blinded to group assignments. No changes in the randomization process occurred during the trial. The study was terminated after the subject assigned as number 177 as recommended by the independent monitoring data report due to high efficacy.

### Procedures

Subjects were to start administration of study drug after randomization. Proxalutamide 200 mg (1 x 200 mg tablets) or matching placebo orally once daily with or without food. Standard of care was offered to all subjects that were not under medical care. Standard of care (treated as per local health care recommendations) consisted of Azithromycin 500mg q.d 5 days, Nitazoxanide 500mg b.i.d 6 days, Dipyrone 1g t.i.d or Paracetamol 750mg t.i.d. (in case of high-grade fever), Ondansetron 8mg t.i.d (in case of nauseas), and Dexamethasone 6mg q.d. (in case of reduction of oxygen saturation above 4p.p. or below 94%).

Treatment compliance was monitored by phone on day 1, 2, 3, 5, and 7, and patients were monitored until complete remission of treatment, and actively searched on days 30, 60, and 90 regarding their health status and persisting or new-onset symptoms.

The following clinical laboratory tests were performed for monitoring the safety profile: hemoglobin (Hb) (g/dL), hematocrit (Ht) (%), neutrophils (/mm^3^), lymphocytes (/mm^3^), monocytes (/mm^3^), eosinophils (/mm^3^), and platelets (*10^3^/mm^3^) (fluorescent flow cytometry and impedance), ALT, AST, total and fractioned bilirubin (mg/dL) (colorimetric assay), blood urea nitrogen (BUN) (mg/dL) (urease with GLDH), calcium (mg/dL) (O-cresolftalein complexone – OCC assay), creatinine (mg/dL) (enzymatic assay), magnesium (mg/dL) (xylidyl blue assay), potassium (mEq/L) (selective electrode assay), sodium (mEq/L) (selective electrode assay), glucose (mg/dL (hexokinase assay), lactate (mmol/L) (enzymatic aasay), ultrasensitive C-Reactive Protein (usCRP) (mg/L) (Latex-intensified immunoturbidimetry assay) and erythrocyte sedimentation rate (ESR) (mm/1h) (capillary photometry assay). The following hormones were performed: total testosterone (ng/dL) (chemiluminescence assay), sex hormone binding globulin (SHBG) (nmol/L) (chemiluminescence assay), dihydrotestosterone (DHT) (pg/mL) (radioimmunoassay), and estradiol (pg/mL) (chemiluminescence assay).

Subjects were treated as outpatients and followed per the normal hospital and clinic protocols for COVID-19 patients. In the event of worsening symptoms patients returned to the hospital and were re-evaluated by the PI. If warranted, patients were admitted to the hospital. Subjects who discontinued treatment for any reason were entered into the long-term follow-up phase of the study. No protocol modifications occurred during the study with female outpatients.

### Study Outcomes

The objective of the study was to evaluate the efficacy of proxalutimide compared to placebo in non-hospitalized COVID-19 female patients. The primary outcome was the hospitalization rate of mild-to-moderate COVID-19 females. To identify hospitalized patients, as secondary outcome, and as the screening method, the COVID-19 8-point ordinal scale was used: 8. Death; 7. Hospitalized, on invasive mechanical ventilation or extracorporealmembrane oxygenation (ECMO); 6. Hospitalized, on high flow oxygen use non-invasive ventilation device; 5. Hospitalized, requiring non-high flow oxygen use; 4. Hospitalized, not requiring supplemental oxygen, requiring ongoing medical care;; 3. Hospitalized, not requiring supplemental oxygen or ongoing medical care; 2. Not hospitalized, with limitation on activities; 1. Not hospitalized, with no limitations on activities. Patients were considered hospitalized if score was three or above at any time during the 30 days after randomization.

### Statistical Methods

Sample size estimate is detailed in Appendix 1. Risk ratio (RR) for hospitalization was used as the measure of efficacy in the analysis of the primary outcome. Kaplan-Meier estimates were used to evaluate the differences of hospitalization rate in the 30-day post-randomization. Intention-to-treat (ITT) analysis was performed. Chi-squared test for independent proportions was employed with statistical significance set at P < 0.05. Stata/SE 17.0 for Mac (StataCorp, TX, USA) was used to perform all statistical analysis.

## Results

### Subjects

One hundred seventy-seven women were randomized. A total of 75 patients in the proxalutamide arm and 102 patients in the placebo arm were followed during the study follow-up period. None of the patients lost to follow-up or discontinued treatment. The CONSORT flowchart of patients enrolled and followed in the study is presented in Figure 1.

**Figure 1.**
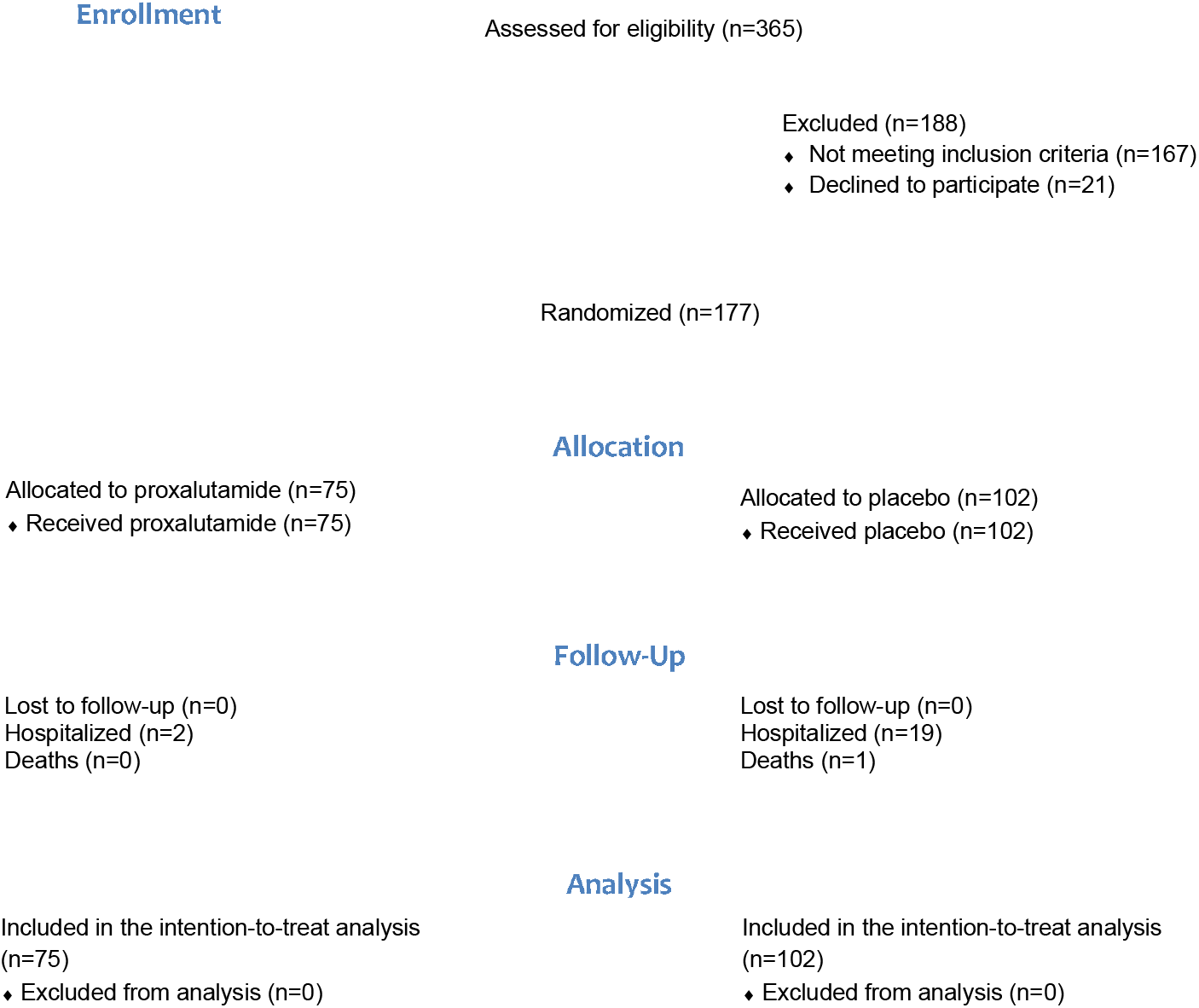
CONSORT Flow Diagram.

Baseline characteristics of both groups are shown in Table 1 and were well similar between proxalutamide and placebo arms. The mean age of subjects was 44.2 years. The study was conducted in Brazil, where the population is ethnically diverse and race mixed. The vast majority of subjects did not self report as being caucasian, black, asian. latin, or indigenous. The use of concomitant medications was difficult to assess due to the heterogeneity of the usual care of outpatients in COVID-19.

**Table 1.** Baseline characteristics of the study arms.

| Characteristics | All<br>(n = 177) | Proxalutamide<br>(n = 75) | Placebo<br>(n = 102) |
| --- | --- | --- | --- |
| Age (yr) mean ± SD | 44.2 ± 12.1 | 45.5 ± 12.6 | 43.2 ± 11.9 |
| **Race** |  |  |  |
| Mixed ethnicity* | 100% (177) | 100% (75) | 100% (102) |
| **No. of coexisting conditions —**<br>**% (Number)** |  |  |  |
| **None** | 57.1% (101) | 50.7% (38) | 61.8% (63) |
| **One** | 27.7% (49) | 26.7% (22) | 26.5% (27) |
| **Two or more** | 15.3% (27) | 20.0% (15) | 11.8% (12) |
| **Coexisting conditions —**<br>**% (Number)** |  |  |  |
| **Type 2 diabetes** | 8.5% (15) | 9.3% (7) | 7.8% (8) |
| **Hypertension** | 31.1% (55) | 24.0% (18) | 20.6% (21) |
| **COPD** | 0.6% (1) | 1.3% (1) | 0 |
| **BMI > 30 kg/m<sup>2</sup>** | 18.1% (32) | 18.7% (14) | 17.6% (18) |

COPD = Chronic Obstructive Pulmonary Disease; BMI = Body mass index;
\* Subjects did not identify as white, mulatto, black, indigenous, latin, or asian

Among the patients randomized to the study, coexisting medical conditions were documented. Variables most likely to affect COVID-19 disease outcomes are reported below for the placebo and treatment arms (Table 3), including hypertension, dyslipidemia, diabetes, pre-diabetes, obesity, asthma, myocardial infarction and COPD.

### Outcomes

During the 30-day period after randomization, statistically significant reduction in the hospitalization rate due to COVID-19 was observed in women taking proxalutamide (two patients; 3%) compared to the placebo (standard of care) (19 patients; 18.6%) (p<0.001) (Table 2). The 30-day hospitalization risk ratio was 0.14 [95% confidence interval (CI), 0.03-0.59], number needed-to-treated (NNT) was 6.3 (95% CI for benefit 3.95-15.12).

**Table 2.** Clinical outcomes throughout 30 days post-randomization.

| **Events**<br>**n (%)** | **Overall**<br>**n = 177** | **Proxalutamide**<br>**n = 75** | **Placebo**<br>**n = 102** | **p-value** |
| --- | --- | --- | --- | --- |
| **Hospitalized** | 21 (11.9%) | 2 (2.7%) | 19 (18.6%) | **0.007** |
| **Not requiring oxygen use** | 2 (1.1%) | 0 (0.0%) | 2 (2.0%) | 0.39 |
| **Requiring non-high flow**<br>**oxygen use** | 19 (10.7%) | 2 (2.7%) | 17 (16.7%) | **0.01** |
| **High-flow oxygen use** | 10 (5.6%) | 0 (0.0%) | 10 (9.8%) | 0.055 |
| **Non-invasive ventilation** | 14 (7.9%) | 0 (0.0%) | 14 (13.7%) | **0.031** |
| **Invasive mechanical**<br>**ventilation** | 5 (2.8%) | 0 (0.0%) | 5 (4.9%) | 0.15 |
| **ECMO** | 0 (0.0%) | 0 (0.0%) | 0 (0.0%) | 1.00 |
| **Vasopressors** | 2 (1.1%) | 0 (0.0%) | 2 (2.0%) | 0.39 |
| **Death** | 1 (0.6%) | 0 (0.0%) | 1 (1.0%) | 0.62 |

ECMO = Extracorporeal membrane oxygenation

Clinical outcomes throughout 30 days post-randomization are presented in Table 2, and the distributions of COVID-19 8-point ordinary scale at baseline, day 15, and day 30 are presented in Table 3. At day 15, none of the patients of the proxalutamide arm and 17 patients of the placebo arm were hospitalized, and in day 30 12 patients in the placebo arm remained hospitalized.

**Table 3.** distributions of COVID-19 8-point ordinary scale at baseline, day 15, and day 30.

| 8-point ordinary scale score [% (n)] | Overall n = 177 | Proxalutamide n = 75 | Placebo n = 102 | p-value |
| --- | --- | --- | --- | --- |
| **Day 0** |  |  |  |  |
| **1** | 70.6% (125) | 68.0% (51) | 72.5% (74) | 0.51 |
| **2** | 29.4% (52) | 32.0% (24) | 27.4% (28) | 0.52 |
| **3** | 0.0% (0) | 0.0% (0) | 0.0% (0) | 1.00 |
| **4** | 0.0% (0) | 0.0% (0) | 0.0% (0) | 1.00 |
| **5** | 0.0% (0) | 0.0% (0) | 0.0% (0) | 1.00 |
| **6** | 0.0% (0) | 0.0% (0) | 0.0% (0) | 1.00 |
| **7** | 0.0% (0) | 0.0% (0) | 0.0% (0) | 1.00 |
| **8** | 0.0% (0) | 0.0% (0) | 0.0% (0) | 1.00 |
| **Day 15** |  |  |  |  |
| **1** | 59.4% (105) | 93.3% (70) | 34.3% (35) | **< 0.0001** |
| **2** | 29.9% (53) | 6.7% (5)* | 47.1% (48)** | **< 0.0001** |
| **3** | 0.6% (1) | 0.0% (0) | 1.0% (1) | 0.62 |
| **4** | 1.1% (2) | 0.0% (0) | 2.0% (2) | 0.39 |
| **5** | 1.1% (2) | 0.0% (0) | 2.0% (2) | 0.39 |
| **6** | 5.1% (9) | 0.0% (0) | 8.8% (9) | 0.07 |
| **7** | 2.8% (5) | 0.0% (0) | 4.9% (5) | 0.15 |
| **8** | 0.0% (0) | 0.0% (0) | 0.0% (0) | 1.00 |
| **Day 30** |  |  |  |  |
| **1** | 77.4% (137) | 98.7% (74) | 61.8% (63) | **0.0008** |
| **2** | 15.8% (28) | 1.3% (1) | 26.5% (27) | **0.003** |
| **3** | 0.6% (1) | 0.0% (0) | 1.0% (1) | 0.62 |
| **4** | 1.1% (2) | 0.0% (0) | 2.0% (2) | 0.39 |
| **5** | 1.7% (3) | 0.0% (0) | 2.9% (3) | 0.27 |
| **6** | 0.6% (1) | 0.0% (0) | 1.0% (1) | 0.62 |
| **7** | 2.8% (5) | 0.0% (0) | 4.9% (5) | 0.15 |
| **8** | 0.0% (0)*** | 0.0% (0) | 0.0% (0)*** | 1.00 |

\*Two hospitalized patients were discharged before day 15.
\*\*Five of the hospitalized patients were discharged before day 15.
\*\*\*Death occurred on day 60.

### Adverse Events

Treatment emergent adverse events (TEAEs) are described in Table 4. TEAEs were reported for 36% of subjects in the proxalutamide arm and for 55% of subjects in the placebo arm. Serious adverse effects (SAEs) were reported for 3% (2/76) of subjects in the proxalutamide arm and for 19% (19/102) of subjects in the placebo arm. Gastrointestinal TEAEs were more frequent in the proxalutamide arm: Diarrhea (17% proxalutamide *versus* 10% placebo), nausea (12% proxalutamide *versus* 6% placebo), abdominal pain (12% proxalutamide versus 5% placebo), and dyspepsia (12% proxalutamide versus 4% placebo).

**Table 4.** Treatment emergent adverse events (TEAEs).

|  | Placebo | Proxalutamide |
| --- | --- | --- |
| **Safety population** | 102 | 75 |
| Number of subjects with 1 or more TEAE | 56 | 27 |
| Number of subjects with SAEs | 19 | 2 |
| *System organ class* |  |  |
| **Gastrointestinal** |  |  |
| Diarrhea | 10 | 13 |
| Nausea | 6 | 9 |
| Abdominal pain | 5 | 9 |
| Vomiting | 0 | 3 |
| Abdominal discomfort | 2 | 6 |
| Dyspepsia | 4 | 9 |
| Heartburn | 3 | 5 |
| **General** |  |  |
| Fatigue | 27 | 4 |
| Fever | 21 | 2 |
| Disease progression | 48 | 8 |
| **Cardiac** |  |  |
| Tachycardia | 6 | 1 |
| **Nervous system** |  |  |
| Headache | 14 | 5 |
| Ageusia | 11 | 3 |
| Anosmia | 15 | 4 |
| **Metabolism and nutrition** |  |  |
| Dehydration | 7 | 8 |
| **Ear and labyrinth** |  |  |
| Ear pain | 7 | 3 |
| **Musculoskeletal and cognitive tissue** |  |  |
| Arthralgia | 14 | 6 |
| Back pain | 13 | 12 |
| Breast pain | 1 | - |
| Pain in extremity | 3 | 10 |
| **Respiratory, thoracic and mediastinal** |  |  |
| Shortness of breath | 28 | 5 |
| **Skin and subcutaneous tissue** |  |  |
| Lesions | 8 | 2 |
| Total TEAE | 251 | 127 |

## DISCUSSION

### Proxalutamide for COVID-19 female outpatients

This study was a randomized, double-blind, placebo controlled, two-arm, parallel study to evaluate hospitalization rates in female COVID-19 outpatients treated with proxalutamide compared to placebo. The study demonstrated a statistically significant and clinically meaningful benefit of proxalutamide over placebo in hospitalization rate (3% vs. 19%, p<0.001).

The robustness and consistency of the positive treatment effect in favor of proxalutamide was confirmed by multiple analysis of intensive care unit (ICU) admission and mechanical ventilation.

The most frequently reported AE (regardless of study drug relation) were disease progression and shortness of breath. The most frequently reported AEs (in>10% or 5%) suspected to be study drug related by patients treated in the proxalutamide group were diarrhea, nausea, abdominal pain, and dyspepsia symptoms. The most frequent symptoms in the placebo group were disease progression and shortness of breath.

Although previous reports demonstrated lack of significant clinical efficacy using nitazoxanide or azithromycin for COVID-19, particularly due to the lower dosage of nitazoxanide used in this study compared to the dosage used in other trials,^22^ whether there were synergistic effects between proxalutamide, nitazoxanide and azithromycin, or whether the benefits could be entirely attributable to proxalutamide, is unelucidated. Also, the number of patients that were treated with nitazoxanide plus azithromycin in each arm is unknown. However, it is clear that proxalutamide made the major role in the prevention of hospitalizations.

### Efficacy of proxalutamide in females versus males

Whether a potent NSAA would be effective for females was uncertain, and based on the translation between the SARS-CoV-2 large dependence on TMPRSS2, indirectly related to androgen activity (*i*.*e*., circulating androgens and sensitivity to androgens) and the presence of infection in a non-neglectable percentage of women, and on previous findings that women with hyperandrogenism experimented more symptoms than non-hyperandrogenic women,^16,17^ while the chronic use of an antiandrogen (spironolactone) mitigated the overrepresentation of symptoms in women.^18,19^

In terms of reduction of hospitalization rate, the efficacy of proxalutamide was demonstrated to be similar between females (86%) and males (91%) if used in the daily dose of 200mg for seven days. Although it was expected that proxalutamide would be effective until certain extent, the fact that the drug has demonstrated similar efficacy between sexes was unexpected, and stimulated us to conduct further randomized clinical trials with proxalutamide in a sex-independent manner. Conversely, apparently proxalutamide nullified the sex-differences on COVID-19, which reinforces that the sex-differences observed in COVID-19 are likely sex steroid-mediated.

### Limitations

Since this is an outpatient study, certainty of the compliance of patients to treatment was a limitation. However, the dropout rate was zero for both arms.

Brazil hospitalization of COVID-19 patients participating in clinical studies is reported by the national health care system. In addition, the standard of care was not consistent. No other limitations have been identified as of the date of February 28, 2020. Criteria for hospitalization was not pre-determined, since it depends on local hospitals protocols. During the period or the study, hospitals were not at full capacity, allowing less severe cases to be hospitalized for preventive reasons, and the decision over hospitalization is not dependent on the principal investigator. Despite the safety profile already preliminarily established, some women declined to participate due to the stigma of use of NSAA for prostate cancer.

### Final discussion

The present study supports that proxalutamide treatment is well tolerated, manageable, and effective in COVID-19 female patients with mild to moderate symptoms. There were no new safety signals or safety concerns in this study.

## CONCLUSIONS

The results of this study indicate that the treatment of COVID-19 outpatients with proxalutamide in combination with standard of care results in clinically meaningful benefit, with significant reduction of hospitalization rate, further supported by lower rate of ICU admission and new oxygen requirement. Treatment with proxalutamide was well tolerated, feasible, and manageable. There were no new safety signals or concerns with proxalutamide treatment.

## Data Availability

Data is available under request for relevant purposes.

